# Altered microbial carbohydrate metabolism is associated with anxiety and gastrointestinal symptoms in patients with Generalized Anxiety Disorder

**DOI:** 10.1101/2025.09.15.25335180

**Authors:** V. Rabbia, M. Amer, G. De Palma, J. Lu, R. Potts, C. James, M. Wolfe, J. Gillard, M. Tersigni, H. Armstrong, SM. Collins, MT. Bailey, M. Surette, R. Anglin, P. Bercik

**Affiliations:** Department of Medicine, Farncombe Family Digestive Health Research Institute, McMaster University, Hamilton, Canada; Department of Psychiatry, McMaster University, Hamilton, Canada; John Buhler Research Centre, University of Manitoba, Winnipeg, Canada; Center for Microbe and Immunity Research, Wexner Research Institute at Nationwide Children’s Hospital, Columbus, OH, USA; Department of Pediatrics, The Ohio State University, College of Medicine, Columbus, OH, USA; National School of Medicine, University of Notre Dame, Fremantle, Australia

**Keywords:** *Microbiota*, *Anxiety*, *Bacteroides*, *Dietary fiber*, *Carbohydrate metabolism*

## Abstract

**Background:** Generalized anxiety disorder (GAD) is a common psychiatric condition, with unknown etiology and pathophysiology. Recent studies have suggested alterations in the microbiota-gut-brain axis may be involved in the development of GAD. We aimed to explore the interactions between the gut microbiota, gastrointestinal and psychiatric symptoms, neuroimmune markers and dietary patterns in patients with GAD.

**Methods:** We recruited 83 GAD patients and 98 age- and sex-matched healthy controls (HC) and assessed their psychiatric and gastrointestinal symptoms, and long-term diet using validated questionnaires. We measured serum and stool neuroimmune markers and metabolites by ELISA and LC-MS, microbiota was analyzed using 16S rRNA gene sequencing with functional predictions by PICRUSt2. Microbial carbohydrate degradation capacity was assessed *ex vivo*. The data was analyzed using classical statistics and machine learning (XGBoost).

**Results:** GAD patients exhibited higher BMI, gastrointestinal symptoms and inflammatory markers, while reporting reduced intake of fiber and other macro- and micronutrients compared to HC. Gastrointestinal symptoms were the most predictive feature separating GAD from HC. GAD patients had a distinct microbiota profile, dominated by *Bacteroides*, compared with a *Prevotella*-dominated microbiota in HC. Carbohydrate degradation pathways were enriched in GAD and strongly associated with Bacteroides abundance. Anxiety scores correlated with *Bacteroides* abundance, carbohydrate degradation pathways and gastrointestinal symptoms, while negatively correlating with dietary fiber intake. *Ex vivo* mucin-to-inulin degradation ratio was higher in GAD and correlated with inflammatory markers.

**Conclusions:** GAD patients exhibited marked gastrointestinal symptoms, elevated immune markers, reduced fiber intake and a *Bacteroides*-dominated microbiota that preferentially degrades mucin. These data suggest that their microbiota adapted to utilize host-derived carbohydrates that may affect the mucus barrier, altering immune homeostasis and leading to gastrointestinal symptoms and anxiety. Dietary interventions, such as gradually increasing fiber intake, could reprogram bacterial carbohydrate metabolism, thus ameliorating gut barrier function and alleviating anxiety and gastrointestinal symptoms.

## Introduction

Generalized anxiety disorder is characterized by excessive and persistent anxiety and worry about different events or activities, which the individual finds difficult to control^1^. Antidepressants are the first choice of treatment for anxiety disorders and have well-documented efficacy in the treatment of GAD. However, it is estimated that around 25 % of patients experience a relapse within 8 years after starting treatment^2^. This limited long-term efficacy likely reflects that these agents act predominantly on specific neurotransmitter systems, whereas the pathophysiology of GAD is multifactorial and not fully understood. Emerging evidence suggests that additional biological factors, including the microbiota, alongside environmental factors, may contribute to disease persistence and are not fully addressed by current therapies

In recent years, accumulating evidence has implicated an altered gut microbiota-brain axis in the pathophysiology of neuropsychiatric disorders, such as major depressive disorder, bipolar disorders, schizophrenia and anxiety^3^. To date, five clinical studies have explored gut microbiota composition in GAD patients, suggesting abnormal bacterial profiles with increased abundance of *Bacteroides*^4–8^. However, these studies did not investigate other putative pathophysiological factors such as diet or microbial metabolic activity, which could provide a better understanding of the impaired gut-brain axis in GAD.

We conducted a comprehensive study in a cohort of well characterized patients and healthy volunteers, investigating gut microbiota profiles together with assessment of long-term dietary patterns, gastrointestinal symptoms, microbial derived metabolites and neuroimmune markers. Our results suggest that patients with GAD have increased inflammatory markers and gastrointestinal symptoms, as well as microbiota dominated with Bacteroides with altered carbohydrate degradation capacity, with preference for mucin.

## Material and Methods

### Clinical cohort

We recruited 83 GAD patients, and 98 age-and-sex matched healthy controls, aged 18 to 65 years from the Anxiety Treatment and Research Centre (St. Joseph’s Health Care, Hamilton, Ontario, Canada). GAD patients fulfilled the Mini International Neuropsychiatric Interview 6.0 GAD diagnosis criteria, while healthy controls had no history of psychiatric disorders. The exclusion criteria for all participants were as follows: (1) antibiotic or probiotic use in the prior 4 weeks; (2) active substance abuse; (3) active eating disorder; (4) pregnancy/lactation; (5) diagnosis of any other axis I or axis II psychiatric illness that was felt to be principal over GAD; (6) diagnosis of any major organic medical illness that may alter the gut microbiota, including gastroenterological, autoimmune, rheumatological or immunological disorders.

From all participants, we obtained demographic data, past medical and psychiatric history, medications, allergies and body mass index (BMI). Participants completed validated questionnaires, including the Depression Anxiety and Stress Scale (DASS), Childhood Trauma Questionnaire, Penn State Worry Questionnaire, Gastrointestinal Symptom Rating Scale (GSRS), short form of Leeds Dyspepsia questionnaire (LDQ), Rome III criteria for irritable bowel syndrome (IBS), and simplified Food Frequency Questionnaire (FFQ; DQES v2, Cancer Council Victoria, Australia). Fecal and blood samples were collected for determination of gut microbiota composition, inflammatory markers, and short-chain fatty acid and neurotransmitter levels.

The study followed the Helsinki Declaration and was approved by the Hamilton Integrated Research Ethics Board on *April 1, 2015 (#14-249)*.

### Sample collection and analysis

Blood samples were collected during the hospital visit at the time of filling up the questionnaires. Stool samples were collected within a week of completing the questionnaires and were delivered to the laboratory at McMaster University within four hours of collection. Stools were aliquoted anaerobically, snap frozen in liquid nitrogen, and stored at −80° C until further use. Subsequent analyses included measurement of inflammatory markers, SCFAs determination, neurotransmitter determination, and microbiota sequencing.

### Assessment of inflammatory markers

Stool human β-defensin-2 and calprotectin were measured according to manufacturer’s instructions via enzyme-linked immunosorbent assay (ELISA) from Immunodiagnostik-K6500 (Bensheim, Germany) and BIOHIT HealthCare Calprotectin ELISA (Ellesmere Port, UK), respectively. Each sample was run in duplicate. Serum CRP was assessed using High Sensitivity CRP ELISA from Life Technologies-KHA0031 (Carlsbad, CA, US), following manufacturer’s instructions.

### Short-chain fatty acid determination

Short-chain fatty acid (acetic, propionic, butyric, isobutyric, isovaleric, pentanoic and lactic acid) analysis was performed at the McMaster Regional Centre of Mass Spectrometry using gas chromatography–mass spectrometry following the protocol by Moreau et al. (2003), with modification for microbiota studies^10,11^.

### Assessment of neurotransmitters

Neurotransmitters from stool samples were quantified by multisegment injection-capillary electrophoresis-mass spectrometry at the Baylor College of Medicine (Houston, USA). All compounds were measured quantitatively, and their identification was confirmed by spiking standards into samples.

### Microbial carbohydrate degradation activity measurement

Minimum culture medium (4 g/L of casamino acids;12.8 g/L of Na_2_HPO_4_ x 7H_2_O; 3 g/L of KH_2_PO_4_; 0.5 g/L of NaCl; 1 g/L of NH_4_Cl; autoclave; 0.5 g/L of L-cysteine (free base); 4 mg/L of thiamine; 1.2 mg/L of hemin; 30 mg/L of histidine; 22 mg/L of MgCl_2_ x 7H_2_O; 0.4 mg/L of FeSO_4_ x 7H_2_O; 1 mg/L of menadione; 8 mg/L of CaCl_2_; 5 µg/L of vitamin B_12_; pH of 7.2) was supplemented with either no carbon source or with 2.4 g/L glucose, 3 g/L inulin (long chain Orafti® IPS, Mannheim, Germany) or 3 g/L mucin from porcine stomach type III (Sigma-Aldrich, St. Louis, USA) (final concentration). Glucose and inulin solutions were filter-sterilized, while mucin was autoclaved separately. Glucose was chosen at 2.4 g/L as 80 % of mucin is made of carbohydrates and it was considered as a positive control for growth. No carbon source solution was used as a negative control. Average of control duplicates with no fecal sample in the 4 different media was used to determine the maximum available carbohydrates concentration.

Fecal samples were diluted 1:10 with saline solution and inoculated into no carbon, glucose, inulin, or mucin fresh media on 96-well plates and incubated at 37° C in anaerobic conditions. After 72 hours, growth was measured at OD_600_ and carbohydrate degradation was determined by the anthrone test. Briefly, sample supernatants were incubated with anthrone solution (2 g/L anthrone in H_2_SO_4_) for 20 minutes at 4° C, 20 minutes at 100° C and 20 minutes at room temperature. Glucose dilutions were included for a standard curve. Anthrone reaction with carbohydrates was measured at OD_620_ in a SpectraMax M3 microplate reader (Molecular Devices, San José, USA) and carbohydrate concentration was determined by interpolating the standard curve. Carbohydrate degradation was calculated as follows:

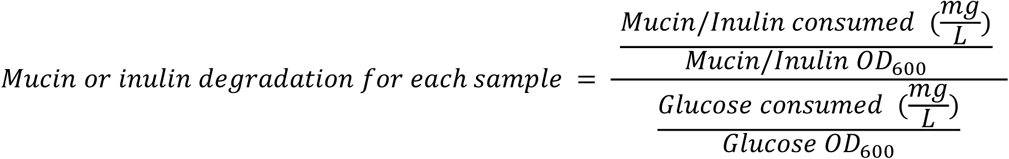

### Microbiota analysis

DNA extraction, amplification and sequencing was carried out following previously described protocols^12,13^. Bacterial 16S rRNA gene profiling was carried out using paired end reads of the V3 region using bar coded Illumina sequencing as described previously^14^, with the modification that bar-codes are included in the forward primer. All PCR products were sequenced on the Illumina MiSeq (San Diego, CA) in the MOBIX-McMaster Genome Center (McMaster University). The microbiota data was processed by an in-house bioinformatics pipeline that included the use of cutadapt^15^, followed dada2^16^. We assigned taxonomy using the Ribosomal Database Project^17^ classifier with the SILVA 138 SSU 2019 training set^18^ with assignment at species level.

We used the amplicon sequence variant (ASV) and taxa tables to run PICRUSt2^19^ for prediction of functional abundances based only on the 16S rRNA gene sequences. We then created a phyloseq object following the phyloseq pipeline^20^. We finally filtered our data, removed all samples with less than 10,000 reads and transformed counts to relative abundance for further analysis. All analyses were conducted in R version 4.1.1 (2021-08-10) using the following packages: dada2, phyloseq, microbiome, vegan, FactoMineR, factoextra, missMDA, XGBoost, ANCOMBC2, dplyr, gtools, ShortRead, phangorn, ape, DECIPHER, graphs were created using ggplot2 and ggpubr.

Alpha diversity measures were calculated using phyloseq objects at ASV level and with raw counts (not relative abundance), while beta diversity Principal Coordinates Analysis (PCoA) plots were generated using the relative abundance transformed phyloseq object collapsed at the desired level and using Bray Curtis distances. AncomBC R package was used to explore microbiota differences between groups.

### Machine learning multivariate analysis

We applied eXtreme Gradient Boosting decision trees (XGBoost) algorithm, generating models to categorize samples into specified output groups (GAD/HC), and subsequently verifying the accuracy of the generated model with a subset of unlabeled samples. The data input included the microbiota data and the metadata, excluding sample IDs, DASS scores, and psychiatric medications, which would emerge as primary classification drivers. We normalized and separated the dataset in 70% training and 30% testing sets. The objective function was set to “binary:logistic”, with 1000 iteration and maximum depth of the tree set to 4.

### Statistical analyses

Chi-square test was used for categorical dependent and independent variable comparison. For continuous dependent variables with 2 groups in the categorical independent variable, a t-test or Mann Whitney was used if the dependent variable was normally or non-normally distributed, respectively. For continuous dependent and independent variables, either Pearson or Spearman correlation was applied if dependent variable was normally or non-normally distributed, respectively. PICRUSt2 data was analyzed by LEfSe (Linear Discriminant Analysis Effect Size) algorithm using the Galaxy web application provided by the Huttenhower Lab^21^. Significant p value was considered as p<0.05 and significant correlation R value was considered as R>0.3. All results were corrected for multiple comparisons, allowing 5% of false discovery rate.

## Results

### GAD patients have higher gastrointestinal symptoms

For general demographics of GAD patients and healthy controls (HC) see Table 1. GAD patients had frequent psychiatric comorbidities, with MDD being the most common diagnosis (57%) (Table 1). Antidepressants and anxiolytics were used by 45% and 18% of GAD patients, respectively. GAD patients reported higher gastrointestinal symptoms compared to HC, in both GSRS (15 vs. 3, p<0.001) and Leeds Dyspepsia (78.5% vs. 28.7%, p<0.001) questionnaires, along with a higher prevalence of IBS diagnosis (36.6% vs. 7.2%, p < 0.001). Additionally, GAD patients exhibited a higher BMI than HC (27.2 vs. 22.9, p=0.002).

**Table 1.** General demographics, psychiatric and gastrointestinal symptoms.

| Demographics |  | GAD<br>(n= 83) | HC<br>(n= 98) | Adjusted<br>p value | Statistical<br>test |
| --- | --- | --- | --- | --- | --- |
| General characteristics | Age | 38.5 (28-46.2) | 32 (22-45) | 0.133 | MW |
|  | Sex (female) | 59 (80.1) | 57 (67.1) | 0.066 | Ch |
|  | Weight (kg) | 74.8 (59.1-82.3) | 67 (59.1-82.3) | 0.078 | MW |
|  | BMI | 27.2 (24-31.6) | 22.9 (21.2-26.7) | 0.002 | MW |
|  | Cigarette smoking | 7 (10.0) | 5 (5.9) | 0.555 | Ch |
|  | Cannabis use | 11 (15.7) | 9 (10.6) | 0.544 | Ch |
|  | Alcohol consumption | 49 (70.0) | 77 (90.6) | 0.007 | Ch |
|  | Vitamin supplements | 24 (29.3) | 28 (29.5) | 1.000 | Ch |
| Medications | Antidepressants | 45 (54.9) | 0 (0) | 2.68x10<sup>-15</sup> | Ch |
|  | Anxiolytics | 18 (22.0) | 0 (0) | 2.71x10<sup>-5</sup> | Ch |
| Comorbid psychiatric disorders | >1 comorbid mental disorder | 50 (61.0) | 0 (0) | 8.27x10<sup>-18</sup> | Ch |
|  | Major depressive disorder | 32 (57.1) | 0 (0) | 1.12x10<sup>-13</sup> | Ch |
|  | Panic disorder | 14 (20) | 0 (0) | 2.78x10<sup>-4</sup> | Ch |
|  | Social anxiety disorder | 8 (14.3) | 0 (0) | 0.005 | Ch |
|  | Posttraumatic stress disorder | 4 (7.1) | 0 (0) | 0.078 | Ch |
|  | Obsessive compulsive disorder | 4 (7.1) | 0 (0) | 0.078 | Ch |
|  | Borderline personality disorder | 10 (22.7) | 0 (0) | 1.33x10<sup>-4</sup> | Ch |
| Psychiatric scores | DASS - Anxiety | 16 (8-20) | 0 (0-2) | 1.18x10<sup>-24</sup> | MW |
|  | DASS - Depression | 14 (6-24) | 0 (0-2) | 1.32x10<sup>-22</sup> | MW |
|  | DASS - Stress | 23 (18-28) | 2 (0-8) | 1.18x10<sup>-24</sup> | MW |
|  | Childhood trauma anxiety | 37 (28-50.8) | 29 (27-33.2) | 1.53x10<sup>-5</sup> | MW |
|  | Penn state worry | 68 (60-73) | 35 (28-41) | 5.57x10<sup>-25</sup> | MW |
| Gastrointestinal scores | IBS-SSS | 30 (36.6) | 7 (7.2) | 2.26x10<sup>-5</sup> | Ch |
|  | GSRS | 15 (8-25) | 3 (0-5) | 9.50x10<sup>-15</sup> | MW |
|  | Leeds dyspepsia | 62 (78.5) | 27 (28.7) | 3.51x10<sup>-7</sup> | Ch |

Values are expressed as number of participants (percentage) for categorical variables, and median (25<sup>th</sup>–75<sup>th</sup> percentile) for continuous variables. Statistical tests: Mann-Whitney U test (MW) and Chi-square test (Ch).

### GAD patients have different dietary patterns with lower fiber intake

Dietary analysis (Table 2) revealed that GAD patients consumed less protein (72.3 vs. 84.7 g/day, p=0.017) and carbohydrates (142.8 vs. 174.0 g/day, p=0.017) compared to HC. Within the carbohydrate category, intake of fiber (15.3 vs. 18.9 g/day, p=0.008) and starch (70.3 vs. 91.4 g/day, p=0.013) was lower in GAD patients. Additionally, intake of several micronutrient were lower in GAD patients, including iron (p=0.005), magnesium (p=0.009), phosphorus (p=0.016), potassium (p=0.02), zinc (p=0.01), niacin (p=0.006), and thiamin (p=0.006).

**Table 2.** Daily dietary patterns Values shown are median (25^th^–75^th^ percentile), representing daily intake.

| Dietary assessment<br>(daily intake) |  | GAD<br>(n= 83) | HC<br>(n= 98) | Adjusted<br>p value |
| --- | --- | --- | --- | --- |
|  | Overall energy (kJ) | 6043 (4845-8088) | 6870 (4845-8088) | 0.052 |
| Macronutrient | Fat (g) | 60.6 (39-81.7) | 63.8 (48.8-82.1) | 0.313 |
|  | Protein (g) | 72.3 (54.4-91) | 84.7(68.2-109.6) | 0.017 |
|  | Carbohydrate (g) | 142.8 (113.8-192.4) | 174.0 (113.8-192.4) | 0.017 |
| Carbohydrate<br>types | Starch (g) | 70.3 (50.7-110.3) | 91.4 (72.3-135.5) | 0.013 |
|  | Sugar (g) | 70.2 (57-90.3) | 83.0 (58.8-100.4) | 0.145 |
|  | Fiber (g) | 15.3 (11.4-21) | 18.9 (15-25.2) | 0.008 |
| Fiber types | Inulin (g) | 0.52 (0.4-0.8) | 0.60 (0.4-0.8) | 0.230 |
|  | Fructooligosaccharide (g) | 1.49 (0.9-2) | 1.43 (1.1-2.4) | 0.201 |
|  | β-glucan (g) | 0.39 (0.2-0.8) | 0.46 (0.3-1.1) | 0.070 |
|  | Pectin (g) | 2.04 (1.2-2.7) | 2.11 (1.2-2.7) | 0.264 |
| Minerals | Calcium (g) | 0.83 (0.5-1) | 0.88 (0.6-1.1) | 0.368 |
|  | Iron (mg) | 8.3 (6.5-11.7) | 10.7 (8.9-13.6) | 0.005 |
|  | Magnesium (mg) | 224.6 (167.3-285.5) | 262.1 (217.7-327.7) | 0.009 |
|  | Phosphorus (g) | 1.26 (0.9-1.5) | 1.45 (1.2-1.8) | 0.016 |
|  | Potassium (g) | 2.3 (1.7-2.6) | 2.5 (2.1-3) | 0.020 |
|  | Sodium (g) | 1.9 (1.3-2.5) | 2.2 (1.7-3) | 0.074 |
|  | Zinc (mg) | 8.6 (6.6-11.4) | 10.6 (8.9-14.2) | 0.010 |
| Vitamins | Vitamin B9 (μg) | 225.3 (139.9-221.5) | 225.3 (177.8-274.1) | 0.007 |
|  | Vitamin A (μg) | 552.5 (386.2-730.6) | 552 (428.9-729.7) | 0.597 |
|  | Vitamin B3 (mg) | 26.22 (19.8-34.9) | 33.38 (19.8-34.9) | 0.006 |
|  | Vitamin B2 (mg) | 1.5 (1.1-1.8) | 1.7 (1.3-2.2) | 0.061 |
|  | Vitamin B1 (mg) | 0.8 (0.6-1.1) | 1.0 (0.8-1.3) | 0.006 |
|  | Vitamin C (mg) | 83.5 (58.5-119.1) | 85.6 (61.5-139.1) | 0.298 |
|  | Vitamin E (mg) | 4.8 (3.5-7.2) | 5.8 (4.2-7.1) | 0.144 |

Values shown are median (25<sup>th</sup>–75<sup>th</sup> percentile), representing daily intake. All statistical comparisons were performed using the Mann-Whitney U test. Folate (Vitamin B9), Retinol equiv. (Vitamin A), Niacin equiv. (Vitamin B3), Riboflavin (Vitamin B2), Thiamin (Vitamin B1).

### GAD patients have higher inflammatory markers and lower SCFA levels

The stool and serum biomarker analysis (Table 3) revealed that GAD patients had higher serum CRP (2.18 mg/L vs. 0.96 mg/L, p=0.039) and stool calprotectin (11.3 mg/kg vs. 0 mg/kg, p=0.02) levels, respectively, compared to HC, while stool beta defensin-2 and serum kynurenine/tryptophan ratio were similar. Total stool SCFA levels were lower in GAD patients compared to HC (45.7 ng/mL vs. 60.9 ng/mL, p=0.002), primarily attributable to reduced acetic acid concentrations (20.4 ng/mL vs. 27.6 ng/mL, p=0.004). No significant differences were found in stool neurotransmitter levels.

**Table 3.** Stool and blood metabolites.

| Stool and blood metabolites |  | GAD<br>(n= 83) | HC<br>(n= 98) | Adjusted<br>p value |
| --- | --- | --- | --- | --- |
| Immune markers<br>(blood) | CRP (mg/L) | 2.18 (1.2-3.2) | 0.96 (1.2-3.2) | 0.039 |
|  | Kynurenine/tryptophan ratio | 38.6 (27.8-53.8) | 39.8 (30.9-49.8) | 0.857 |
| Immune markers<br>(stool) | Calprotectin (mg/Kg) | 11.3 (1.8-39.7) | 0 (0-26.9) | 0.020 |
|  | Beta defensin-2 (ng/mL) | 32.6 (19.5-77.2) | 30.1 (12.6-53) | 0.294 |
| Short-chain<br>fatty acids (SCFA)<br>(stool) | All SCFAs (ng/mL) | 45.7 (28.4-58.8) | 60.9 (36.8-84.2) | 0.002 |
|  | Acetic acid (ng/mL) | 20.4 (13.6-30.6) | 27.6 (21-37.9) | 0.004 |
|  | Propionic acid (ng/mL) | 9.0 (4.8-12.7) | 11.3 (7.4-14.5) | 0.065 |
|  | Butyric acid (ng/mL) | 9.3 (5.5-14) | 11.5 (6.4-21) | 0.064 |
| Neurotransmitter<br>related compounds<br>(stool) | GABA (ng/mL) | 87.9 (57.6-252.9) | 137.2 (82.6-389.1) | 0.097 |
|  | Glutamate (μg/mL) | 29.6 (2.2-5.5) | 38.9 (25-51.2) | 0.265 |
|  | Glutamine (μg/mL) | 4.9 (2.7-6.5) | 3.7 (2.3-5.6) | 0.203 |
|  | Dopamine (ng/mL) | 10 (0-27.9) | 15.0 (0-60.5) | 0.465 |
|  | Serotonin (ng/mL) | 34.5 (1.9-73.2) | 40.8 (23.4-71.4) | 0.857 |
|  | 5-hydroxyindoleacetic acid<br>(ng/mL) | 94.8 (15.1-164.2) | 114.1 (35.9-194.8) | 0.368 |
|  | Tryptophan (μg/mL) | 14.8 (10.2-21.3) | 12.1 (8.8-20) | 0.545 |
|  | 5-Hydroxytryptophan (ng/mL) | 5.2 (2.6-11) | 6.5 (2.2-21.2) | 0.555 |
|  | N-Acetyl-serotonin (ng/mL) | 1.2 (0.6-3) | 2.2 (1-3.2) | 0.224 |

Values are expressed as median (25<sup>th</sup>–75<sup>th</sup> percentile). Mann-Whitney U test was used for all comparisons.

### Microbiota profiles differ between GAD and HC

There were no differences in alpha diversity indices, including Chao1 (Figure 1A), Shannon, Simpson, Inverse Simpson, and Berger-Parker dominance (Mann-Whitney U test; p=0.21, p=0.18, p=0.7, p=0.7, and p=0.94, respectively). However, beta diversity differed when assessed by Bray-Curtis distance (Figure 1B; Adonis PERMANOVA, R^2^=0.033; p=0.003), as well as UniFrac, weighted UniFrac and Jaccard distance for PCoA plot construction (not shown). Despite this, the explained variability (R^2^) between groups was very low, suggesting that other variables are likely contributing to the heterogeneity of the data.

**Figure 1.**
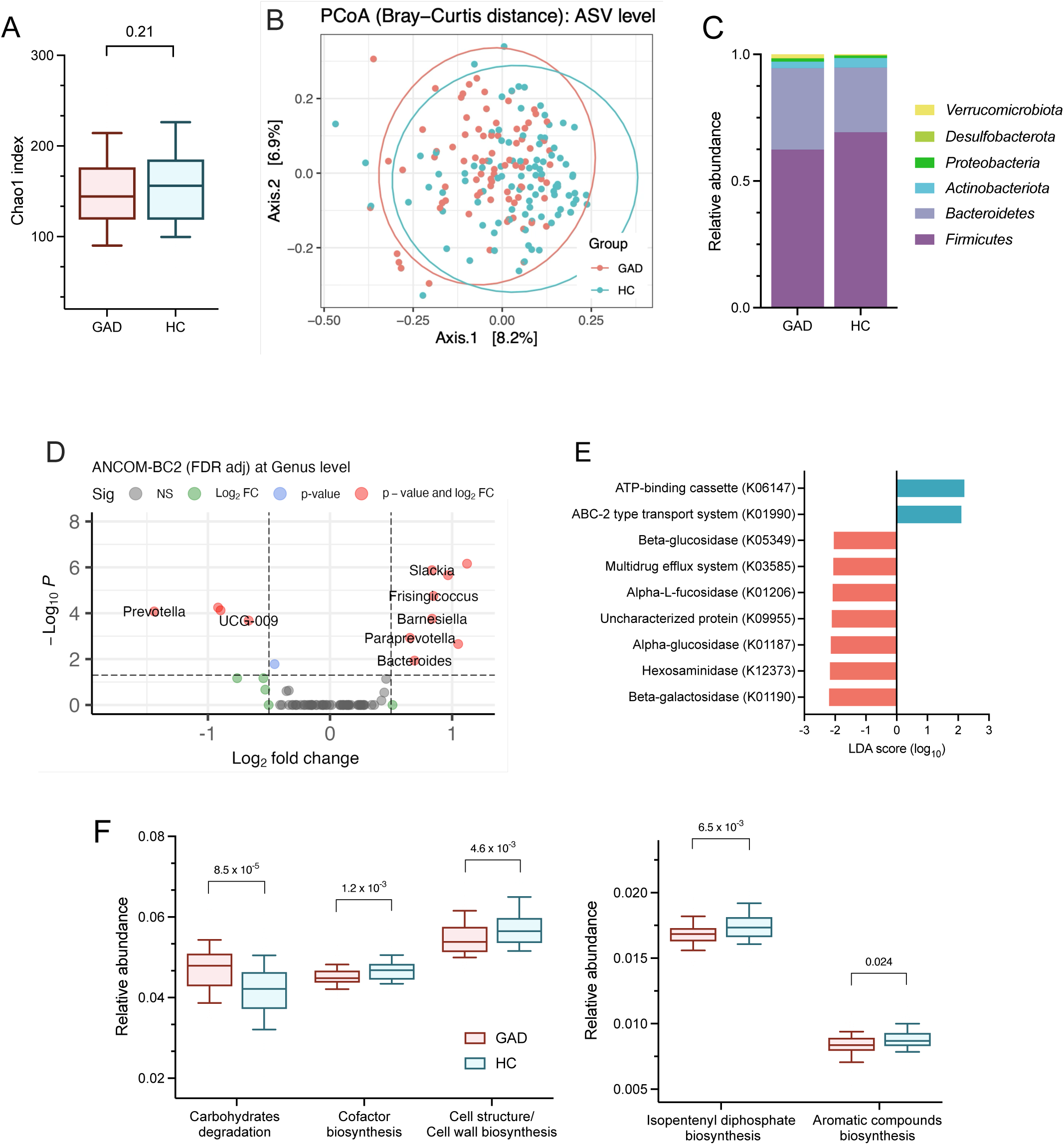
Gut microbiota composition and predicted function differ between GAD patients and HC. A) Alpha diversity measured by Chao1 index. Significance was determined by Mann– Whitney U test. Data presented as median ± interquartile range. B) Beta diversity assessed using Bray–Curtis dissimilarity. Group differences tested using PERMANOVA, p = 0.003. C) Relative abundance of bacterial taxa at the phylum level. D) Volcano plot of ANCOMBC2 genus-level differential abundance between GAD and HC. Significance threshold was set at adjusted p < 0.05 and log₂FC > 0.5. E) Differential abundance of KEGG Orthology identified with PICRUSt2 and analyzed by LEfSe (LDA score > 2.2, p < 0.05). HC was set as reference group. E) Predicted functional pathway super classes identified with PICRUSt2. Statistical comparisons performed using the Mann–Whitney U test.

Further examination at phylum and genus level revealed significant group differences (Figure 1C). GAD patients showed higher levels of *Bacteroidetes* and *Actinobacteria*, and lower levels of *Firmicutes* compared to HC (Mann-Whitney U test; p=0.01, p=0.03, and p=0.01, respectively). We performed ANCOMBC2 at genus level and found that several taxa were differentially abundant in the groups, including increased *Slackia, Barnesiella* and *Bacteroides*, and decreased *Prevotella* in GAD patients (Figure 1D).

### GAD patients have higher predicted microbial carbohydrate degradation

Predicted metagenomic function analysis identified 7,094 KEGG ortholog genes across 413 MetaCyc pathways. Most of the enzymes higher in the microbiota of GAD patients were linked to multiple carbohydrate degradation pathways (K01190, K12373, K05349, K01187, and K01206; Figure 1E) including glucosidases, fucosidase, hexosaminidase and galactosidase enzymes. By collapsing these pathways into their respective super classes using the published classification^21^, we found that the most significant difference between groups was higher carbohydrate degradation, followed by cofactor biosynthesis and cell wall biosynthesis (Figure 1F).

### Gastrointestinal symptoms are the strongest predictor of GAD

To explore relationships between GAD and clinical, microbial, and dietary variables, we employed a supervised machine learning analysis using the XGBoost algorithm. The model demonstrated strong predictive power in classifying GAD and HC group with an accuracy of 87.7%. Gastrointestinal symptoms as a group were the top predictors distinguishing GAD from HC, and within this group, abdominal pain was the dominant feature driving the model’s classification (Figure 2A), suggesting a strong role of the gut-brain axis in GAD.

**Figure 2.**
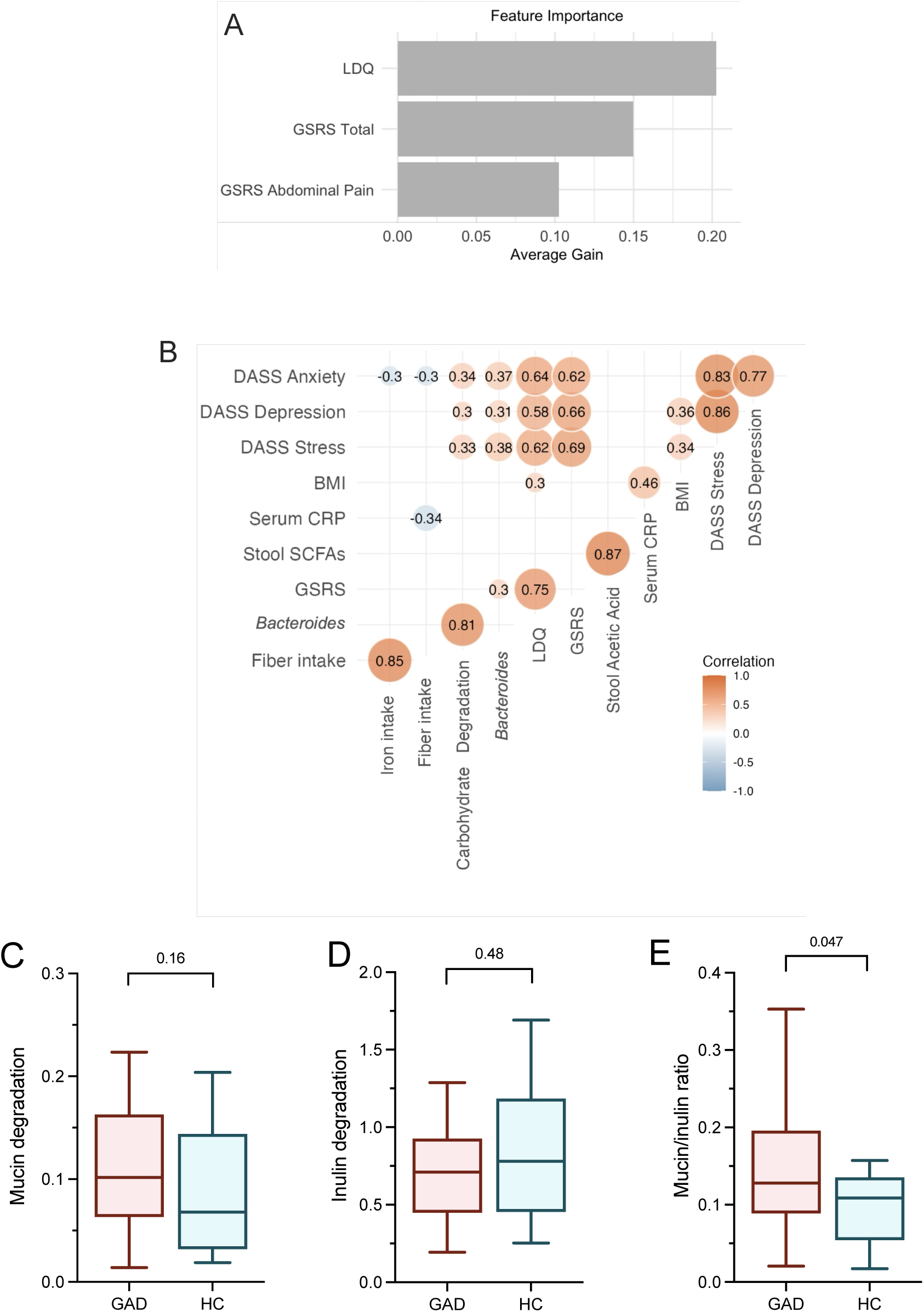
Host and microbial predictors of GAD, including shifts in carbohydrate degradation capacity. A) Feature importance ranking plot from XGBoost model classifying GAD vs. HC, highlighting the most predictive variable. B) Correlation matrix showing relationships between mental health scores (DASS), gastrointestinal symptom scores (LDQ, GSRS), microbial taxa and functions, diet, and host biomarkers. Only correlations with p < 0.05 and |R| > 0.3 are shown. C-E) *In vitro* carbohydrate degradation capacity of fecal microbiota for inulin (C), mucin (D), and the mucin-to-inulin degradation ratio (E). Statistical significance was determined using the Mann–Whitney U test. Data are presented as median ± interquartile range.

### Anxiety correlates with abdominal pain, fiber intake, and bacterial carbohydrate degradation

To better understand the relationships between the individual variables in this study, we constructed a multiple correlation plot, including only significant correlations (p<0.05) with R>0.3 (Figure 2B, Figure 2S). Supporting the results obtained by XGBoost, DASS anxiety scores strongly correlated with the severity of gastrointestinal symptoms, as assessed by the GSRS (R=0.62) and Leeds Dyspepsia (R=0.64) questionnaires. A detailed analysis revealed that abdominal pain was the GSRS subdomain most closely linked to anxiety scores (R=0.59; Figure S1).

Anxiety scores negatively correlated with both fiber intake (R=-0.30) and iron intake (R=-0.30). Upon further exploration, inulin (R=-0.21) and fructooligosaccharides (FOS; R=-0.22) were identified as the specific fibers that were most strongly inversely correlated with anxiety scores (Figure S1). Further, fiber intake negatively correlated with (R= −0.34) with serum CRP.

Anxiety scores positively correlated with *Bacteroides* abundance (R=0.37) and carbohydrate degradation pathways (R=0.34), which were also strongly correlated to each other (R=0.81), suggesting that *Bacteroides* abundance may play a significant role in gut carbohydrate metabolism. To further explore this relationship, we investigated the *ex vivo* carbohydrate degradation activity of the stool samples, focusing on dietary carbohydrates, specifically inulin (the fiber type that showed the strongest negative correlation to anxiety scores in GAD patients), and host-derived carbohydrates, specifically mucin.

### GAD microbiota has an altered carbohydrate utilization dynamics

The ability to degrade inulin and mucin was assessed in a randomly selected subset of stool microbiota samples from our cohort (41 HC and 28 GAD patients). Although there was a trend toward increased mucin degradation and reduced inulin degradation in GAD samples, neither reached statistical significance (Figure 2C, D). However, mucin-to-inulin ratio was significantly increased in GAD samples (Figure 2E), suggesting a shift in microbial carbohydrate utilization dynamics. This may reflect a relative increase in reliance on host-derived carbohydrates by the microbiota in GAD, potentially driven by reduced availability of dietary fibers. Additionally, mucin-to-inulin degradation ratio strongly correlated to serum CRP (R=0.91, p=0.004) and borderline correlated to anxiety scores (R=0.4, p=0.06) in GAD patients.

## Discussion

The role of the gut microbiota in the pathophysiology of GAD has been explored in recent years, revealing altered microbiota profiles, but the underlying mechanisms remain unclear. In this study, we conducted a comprehensive characterization of gut microbiota composition, predicted microbial metabolic functions, dietary patterns, and gastrointestinal and psychiatric symptoms in a well-defined cohort of GAD patients and healthy controls. Our main findings involve a GAD cohort marked by: (i) elevated gastrointestinal symptoms, particularly abdominal pain; (ii) reduced intake of several micronutrients, including dietary fiber; (iii) a distinct microbiota composition marked by *Bacteroides* dominance; (iv) lower levels of fecal SCFAs; and (v) enrichment in microbial carbohydrate degradation pathways predicted by PICRUSt2. Several of these variables, including gastrointestinal symptoms, *Bacteroides* abundance, and dietary fiber intake, were significantly correlated with anxiety severity. Our findings support the microbiota-gut-brain axis as a critical mediator of anxiety symptoms, highlighting the interconnected roles of diet, gastrointestinal distress, and immune activation.

Previous studies have consistently reported alterations in the gut microbiota of GAD patients, particularly increased *Bacteroides* abundance and reduced alpha diversity ^4–8^. However, most of these studies focused on taxonomic profiles and did not explore functional microbial features or host factors such as diet, biomarkers or gastrointestinal symptoms. Our findings confirm and extend previous reports by integrating multi-dimensional data and supporting the importance of host-microbiota-diet interactions in GAD. Notably, a *Bacteroides*-dominant microbiota has been linked to Western, low-fiber diets^22–24^, and this pattern was predominant in our GAD patients. In contrast, healthy controls displayed a *Prevotella*-dominant profile with higher reported fiber intake, consistent with associations between *Prevotella* and healthier lifestyles, including higher fiber intake and regular exercise^22,25,26^.

Reduced fiber intake in our GAD cohort was accompanied by lower levels of fecal SCFAs, especially acetate. These results are in line with prior evidence indicating that fiber-rich diets support SCFA production and maintain microbial diversity ^27^. SCFAs such as acetate and butyrate play crucial roles in gut physiology, modulating inflammation and affecting brain function^28^. Moreover, preclinical studies have demonstrated that SCFA supplementation can reduce anxiety-like behavior in animal models, suggesting a potential link between SCFAs and mood regulation in the context of GAD^29–31^.

Predicted metagenomic analysis using PICRUSt2 revealed enrichment of carbohydrate-degrading enzymes in the microbiota of GAD patients. These enzymes include glycosidases such as K01190, K12373, K05349, K01187, and K01206, involved in the degradation of mucin-type glycans and complex polysaccharides, several of which have been implicated in adaptive responses to fiber restriction^32^. A similar metabolic adaptation was observed in our GAD microbiota and was strongly correlated with *Bacteroides* relative abundance. This genus is known for its broad metabolic capacity and ability to switch rapidly between dietary and host-derived carbohydrates ^33,34^. *In vitro* studies have shown that *Bacteroides uniformis* upregulates mucin-degrading enzymes when grown on mucin, including those identified in our study, while downregulating sugar transporters like K01990 when grown on inulin^35^. These findings suggest that the GAD microbiota may be metabolically more versatile and better equipped to utilize a broader range of carbohydrates, including host-derived glycans, especially under low fiber conditions or other disease-associated factors.

To complement our predictions, we assessed *ex vivo* degradation of mucin and inulin in a subset of stool samples. Although degradation of individual substrates did not significantly differ between groups, the mucin-to-inulin degradation ratio was significantly elevated in GAD. This observation may reflect relatively altered carbohydrate utilization dynamics in GAD-associated microbiota when compared to HC. The intestinal epithelium constitutively secretes mucin to renew the mucus layer, and together with gut bacteria that metabolize mucin glycans, contributes to its constant turnover in a healthy gut; however, excessive degradation of the mucus layer could result in increased intestinal permeability and subsequent immune activation^36–38^. Further, a recent study found that expansion of *Bacteroides thetaiotaomicron* causes mucosal barrier damage and immune activation, which can be reversed by supplementing xylose (fiber-derived monosaccharide) that downregulates the expression of mucin-degrading enzymes^39^. In line with this hypothesis, we found that the mucin-to-inulin degradation ratio was strongly associated with serum CRP levels. Our data suggest that dietary factors may influence microbial enzymatic activity in a clinically relevant manner.

Importantly, gastrointestinal symptoms emerged as the strongest predictor of GAD status in our machine learning model, surpassing microbial or dietary variables. This finding reinforces previous work demonstrating the high burden of GI complaints in anxiety disorders ^40–42^, and supports the relevance of gut-focused symptomatology in GAD. In our cohort, immune activation was also evident, as shown by elevated levels of serum CRP and fecal calprotectin, aligning with prior studies reporting increased TNF-α and IFN-γ in GAD^43–45^. Our findings suggest that microbial carbohydrate metabolism, shaped by both dietary intake and microbiota composition, may be a key factor underlying gastrointestinal symptoms in GAD, and thus a viable target for intervention.

Dietary fiber, particularly inulin and FOS, emerges as a simple and non-invasive intervention strategy to support gut health through offering an alternative energy source to host glycans to gut microbiota, potentially decreasing anxiety symptoms in GAD patients. Previous studies support this idea by linking anxiety and depression to low fiber^46–48^ and Western diets in general^49,50^. Moreover, several studies have found a beneficial effect of FOS administration on anxiety scores in patients with IBS^51–54^.

In conclusion, our study identifies a distinct clinical and microbial profile in GAD, characterized by reduced fiber intake, elevated gastrointestinal symptoms, increased abundance of *Bacteroides*, lower SCFA levels, and enrichment in carbohydrate-degrading microbial pathways. These interconnected findings support the idea that dietary patterns shape microbiota structure and function in ways that may influence both gut and mental health. Our results point to microbial carbohydrate metabolism as a potentially important contributor to anxiety symptoms in GAD patients. Increasing fermentable fiber intake, particularly in individuals with low dietary fiber and significant gastrointestinal burden, should be further explored as a simple strategy to modulate microbiota activity, alleviate symptoms and improve clinical outcomes in GAD.

## Data Availability

All data produced in the present study are available upon reasonable request to the authors

---

**Figure S1.**
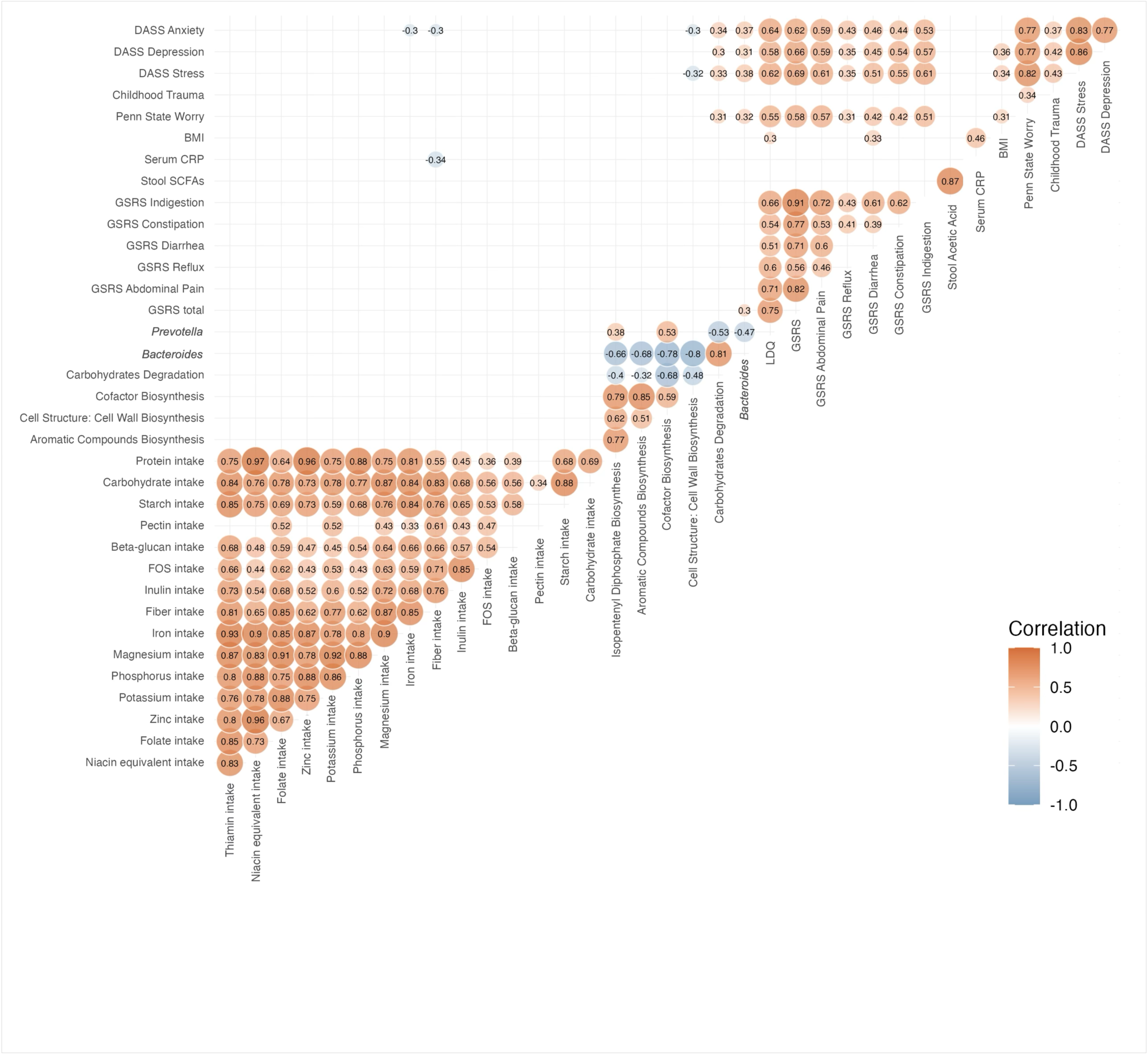
Correlation of clinical, microbial and dietary factors. Complete correlation matrix including mental health test scores (DASS), gastrointestinal symptom scores (LDQ, GSRS), dietary factors, microbial taxa and predicted functions, and host biomarkers. Only correlations with p < 0.05 and |R| > 0.3 are displayed.

## References

1. Diagnostic and Statistical Manual of Mental Disorders : DSM-5-TR. 5th edition, text. American Psychiatric Association Publishing; 2022.

2. Francis JL, Moitra E, Dyck I, Keller MB. The impact of stressful life events on relapse of generalized anxiety disorder. Depress Anxiety. 2012;29(5):386–391. 10.1002/da.20919

3. Iannone LF, Preda A, Blottière HM, et al. Microbiota-gut brain axis involvement in neuropsychiatric disorders. Expert Rev Neurother. 2019;19(10):1037–1050. doi:10.1080/14737175.2019.1638763

4. Jiang HY, Zhang X, Yu ZH, et al. Altered gut microbiota profile in patients with generalized anxiety disorder. J Psychiatr Res. 2018;104:130–136. doi:10.1016/j.jpsychires.2018.07.007

5. Chen YH, Bai J, Wu D, et al. Association between fecal microbiota and generalized anxiety disorder: Severity and early treatment response. J Affect Disord. 2019;259:56–66. 10.1016/j.jad.2019.08.014

6. Mason BL, Li Q, Minhajuddin A, et al. Reduced anti-inflammatory gut microbiota are associated with depression and anhedonia. J Affect Disord. 2020;266(October 2019):394–401. doi:10.1016/j.jad.2020.01.137

7. Guo X, Lin F, Yang F, Chen J, Cai W, Zou T. Gut microbiome characteristics of comorbid generalized anxiety disorder and functional gastrointestinal disease: Correlation with alexithymia and personality traits. Front Psychiatry. 2022;13. doi:10.3389/fpsyt.2022.946808

8. Dong Z, Shen X, Hao Y, et al. Gut Microbiome: A Potential Indicator for Differential Diagnosis of Major Depressive Disorder and General Anxiety Disorder. Front Psychiatry. 2021;12. doi:10.3389/fpsyt.2021.651536

9. Moreau NM, Goupry SM, Antignac JP, et al. Simultaneous measurement of plasma concentrations and 13C-enrichment of short-chain fatty acids, lactic acid and ketone bodies by gas chromatography coupled to mass spectrometry. J Chromatogr B Analyt Technol Biomed Life Sci. 2003;784(2):395–403. doi:10.1016/S1570-0232(02)00827-9

10. Samuel BS, Gordon JI. A Humanized Gnotobiotic Mouse Model of Host-Archaeal-Bacterial Mutualism. Proceedings of the National Academy of Sciences - PNAS. 2006;103(26):10011–10016. doi:10.1073/pnas.0602187103

11. Gordon JI, Turnbaugh PJ, Ley RE, Mahowald MA, Magrini V, Mardis ER. An obesity-associated gut microbiome with increased capacity for energy harvest. Nature. 2006;444(7122):1027–1131. doi:10.1038/nature05414

12. Sibley CD, Grinwis ME, Field TR, et al. Culture enriched molecular profiling of the cystic fibrosis airway microbiome. PLoS One. 2011;6(7):e22702–e22702. doi:10.1371/journal.pone.0022702

13. Whelan FJ, Verschoor CP, Stearns JC, et al. The loss of topography in the microbial communities of the upper respiratory tract in the elderly. Ann Am Thorac Soc. 2014;11(4):513–521. doi:10.1513/AnnalsATS.201310-351OC

14. Bartram AK, Lynch MDJ, Stearns JC, Moreno-Hagelsieb G, Neufeld JD. Generation of Multimillion-Sequence 16S rRNA Gene Libraries from Complex Microbial Communities by Assembling Paired-End Illumina Reads. Appl Environ Microbiol. 2011;77(11). doi:10.1128/AEM.02772-10

15. Saeidipour B, Bakhshi S. The relationship between organizational culture and knowledge management,& their simultaneous effects on customer relation management. Adv Environ Biol. 2013;7(10):2803–2809.

16. Callahan BJ, McMurdie PJ, Rosen MJ, Han AW, Johnson AJA, Holmes SP. DADA2: High-resolution sample inference from Illumina amplicon data. Nat Methods. 2016;13(7). doi:10.1038/nmeth.3869

17. Wang Q, Garrity GM, Tiedje JM, Cole JR. Naïve Bayesian Classifier for Rapid Assignment of rRNA Sequences into the New Bacterial Taxonomy. Appl Environ Microbiol. 2007;73(16). doi:10.1128/AEM.00062-07

18. DeSantis TZ, Hugenholtz P, Larsen N, et al. Greengenes, a Chimera-Checked 16S rRNA Gene Database and Workbench Compatible with ARB. Appl Environ Microbiol. 2006;72(7). doi:10.1128/AEM.03006-05

19. Langille MGI, Zaneveld J, Caporaso JG, et al. Predictive functional profiling of microbial communities using 16S rRNA marker gene sequences. Nat Biotechnol. 2013;31(9). doi:10.1038/nbt.2676

20. McMurdie PJ, Holmes S. phyloseq: An R Package for Reproducible Interactive Analysis and Graphics of Microbiome Census Data. PLoS One. 2013;8(4). doi:10.1371/journal.pone.0061217

21. Segata N, Izard J, Waldron L, et al. Metagenomic biomarker discovery and explanation. Genome Biol. 2011;12(6). doi:10.1186/gb-2011-12-6-r60

22. Wang J, Li W, Wang C, et al. Enterotype *Bacteroides* Is Associated with a High Risk in Patients with Diabetes: A Pilot Study. Peterson JM, ed. J Diabetes Res. 2020;2020:6047145. doi:10.1155/2020/6047145

23. Arumugam M, Raes J, Pelletier E, et al. Enterotypes of the human gut microbiome. Nature. 2011;473(7346):174–180. doi:10.1038/nature09944

24. Zhang F, Fan D, Huang J lin, Zuo T. The gut microbiome: linking dietary fiber to inflammatory diseases. Medicine in Microecology. 2022;14:100070. 10.1016/j.medmic.2022.100070

25. Al Bataineh MT, Dash NR, Bel Lassen P, et al. Revealing links between gut microbiome and its fungal community in Type 2 Diabetes Mellitus among Emirati subjects: A pilot study. Sci Rep. 2020;10(1):9624. doi:10.1038/s41598-020-66598-2

26. Petersen LM, Bautista EJ, Nguyen H, et al. Community characteristics of the gut microbiomes of competitive cyclists. Microbiome. 2017;5(1):1–13. doi:10.1186/s40168-017-0320-4

27. Chen T, Long W, Zhang C, Liu S, Zhao L, Hamaker BR. Fiber-utilizing capacity varies in Prevotella-versus Bacteroides-dominated gut microbiota. Sci Rep. 2017;7(1):1–7. doi:10.1038/s41598-017-02995-4

28. Martin-Gallausiaux C, Marinelli L, Blottière HM, Larraufie P, Lapaque N. SCFA: mechanisms and functional importance in the gut. Proceedings of the Nutrition Society. 2021;80(1):37–49. doi:10.1017/S0029665120006916

29. Palepu MSK, Gajula SNR, Malleshwari K, Sonti R, Dandekar MP. SCFAs Supplementation Rescues Anxiety- and Depression-like Phenotypes Generated by Fecal Engraftment of Treatment-Resistant Depression Rats. ACS Chem Neurosci. 2024;15(5):1010–1025. doi:10.1021/acschemneuro.3c00727

30. van de Wouw M, Boehme M, Lyte JM, et al. Short-chain fatty acids: microbial metabolites that alleviate stress-induced brain–gut axis alterations. J Physiol. 2018;596(20):4923–4944. 10.1113/JP276431

31. Wu JT, Sun CL, Lai TT, et al. Oral short-chain fatty acids administration regulates innate anxiety in adult microbiome-depleted mice. Neuropharmacology. 2022;214. doi:10.1016/j.neuropharm.2022.109140

32. Desai MS, Seekatz AM, Koropatkin NM, et al. A Dietary Fiber-Deprived Gut Microbiota Degrades the Colonic Mucus Barrier and Enhances Pathogen Susceptibility. Cell. 2016;167(5):1339–1353.e21. doi:10.1016/j.cell.2016.10.043

33. Zafar H, Saier MHJ. Gut Bacteroides species in health and disease. Gut Microbes. 2021;13(1):1–20. doi:10.1080/19490976.2020.1848158

34. Raimondi S, Musmeci E, Candeliere F, Amaretti A, Rossi M. Identification of mucin degraders of the human gut microbiota. Sci Rep. 2021;11(1):11094. doi:10.1038/s41598-021-90553-4

35. Benítez-Páez A, Gómez Del Pulgar EM, Sanz Y. The Glycolytic Versatility of Bacteroides uniformis CECT 7771 and Its Genome Response to Oligo and Polysaccharides. Front Cell Infect Microbiol. 2017;7:383. doi:10.3389/fcimb.2017.00383

36. Schroeder BO, Birchenough GMH, Ståhlman M, et al. Bifidobacteria or Fiber Protects against Diet-Induced Microbiota-Mediated Colonic Mucus Deterioration. Cell Host Microbe. 2018;23(1):27–40.e7. doi:10.1016/j.chom.2017.11.004

37. Cani PD, Neyrinck AM, Fava F, et al. Selective increases of bifidobacteria in gut microflora improve high-fat-diet-induced diabetes in mice through a mechanism associated with endotoxaemia. Diabetologia. 2007;50(11):2374–2383. doi:10.1007/s00125-007-0791-0

38. Cani PD, Bibiloni R, Knauf C, et al. Changes in Gut Microbiota Control Metabolic Endotoxemia-Induced Inflammation in High-Fat Diet–Induced Obesity and Diabetes in Mice. Diabetes. 2008;57(6):1470–1481. doi:10.2337/db07-1403

39. Hayase E, Hayase T, Jamal MA, et al. Mucus-degrading Bacteroides link carbapenems to aggravated graft-versus-host disease. Cell. 2022;185(20):3705–3719.e14. 10.1016/j.cell.2022.09.007

40. Mussell M, Kroenke K, Spitzer RL, Williams JBW, Herzog W, Löwe B. Gastrointestinal symptoms in primary care: Prevalence and association with depression and anxiety. J Psychosom Res. 2008;64(6):605–612. doi:10.1016/j.jpsychores.2008.02.019

41. Söderquist F, Syk M, Just D, et al. A cross-sectional study of gastrointestinal symptoms, depressive symptoms and trait anxiety in young adults. BMC Psychiatry. 2020;20(1). doi:10.1186/s12888-020-02940-2

42. Cobos-Palacios L, Ruiz-Moreno MI, Vilches-Perez A, et al. Metabolically healthy obesity: Inflammatory biomarkers and adipokines in elderly population. PLoS One. 2022;17(6 June). doi:10.1371/journal.pone.0265362

43. Costello H, Gould RL, Abrol E, Howard R. Systematic review and meta-analysis of the association between peripheral inflammatory cytokines and generalised anxiety disorder. BMJ Open. 2019;9(7):e027925. doi:10.1136/bmjopen-2018-027925

44. Tang Z, Ye G, Chen X, et al. Peripheral proinflammatory cytokines in Chinese patients with generalised anxiety disorder. J Affect Disord. 2018;225:593–598. doi:10.1016/j.jad.2017.08.082

45. Hou R, Garner M, Holmes C, et al. Peripheral inflammatory cytokines and immune balance in Generalised Anxiety Disorder: case-controlled study. Brain Behav Immun. 2017;62:212–218. doi:10.1016/j.bbi.2017.01.021

46. Taylor AM, Holscher HD. A review of dietary and microbial connections to depression, anxiety, and stress. Nutr Neurosci. 2020;23(3):237–250. doi:10.1080/1028415X.2018.1493808

47. Bloch M, Rubinow DR, Schmidt PJ, Lotsikas A, Chrousos GP, Cizza G. Cortisol response to ovine corticotropin-releasing hormone in a model of pregnancy and parturition in euthymic women with and without a history of postpartum depression. Journal of Clinical Endocrinology and Metabolism. 2005;90(2):695–699. doi:10.1210/jc.2004-1388

48. Kris-Etherton PM, Petersen KS, Hibbeln JR, et al. Nutrition and behavioral health disorders: Depression and anxiety. Nutr Rev. 2021;79(3):247–260. doi:10.1093/nutrit/nuaa025

49. Jacka FN, Pasco JA, Mykletun A, et al. Association of Western and Traditional Diets With Depression and Anxiety in Women. American Journal of Psychiatry. 2010;167(3):305–311. doi:10.1176/appi.ajp.2009.09060881

50. Sadeghi O, Keshteli AH, Afshar H, Esmaillzadeh A, Adibi P. Adherence to Mediterranean dietary pattern is inversely associated with depression, anxiety and psychological distress. Nutr Neurosci. 2021;24(4):248–259. doi:10.1080/1028415X.2019.1620425

51. Silk DBA, Davis A, Vulevic J, Tzortzis G, Gibson GR. Clinical trial: the effects of a trans-galactooligosaccharide prebiotic on faecal microbiota and symptoms in irritable bowel syndrome. Aliment Pharmacol Ther. 2009;29(5):508–518. doi:10.1111/j.1365-2036.2008.03911.x

52. Azpiroz F, Dubray C, Bernalier-Donadille A, et al. Effects of scFOS on the composition of fecal microbiota and anxiety in patients with irritable bowel syndrome: a randomized, double blind, placebo controlled study. Neurogastroenterology and motility. 2017;29(2):e12911-n/a. doi:10.1111/nmo.12911

53. Schmidt K, Cowen PJ, Harmer CJ, Tzortzis G, Errington S, Burnet PWJ. Prebiotic intake reduces the waking cortisol response and alters emotional bias in healthy volunteers. Psychopharmacology (Berl). 2015;232(10):1793–1801. doi:10.1007/s00213-014-3810-0

54. Herkes S, Lukaszuk J, Walker D, Shokrani M. Inulin, Containing Fructo-oligosaccharides, and the Generalized Anxiety Disorder 7-Item Scale Scores in College Students. J Acad Nutr Diet. 2022;122(10):A106–A106. doi:10.1016/j.jand.2022.08.057

